# Surgical Outcomes of Myelomeningocele Repair: a 20-Year Experience from a Single Center in a Middle-Income Country

**DOI:** 10.1101/2021.11.07.21266030

**Authors:** Sina Zoghi, Maryam Feili, Mohammad Amin Mosayebi, Mohammad Amin Afifi, Afrooz Feili, Mohammad Sadegh Masoudi, Reza Taheri

**Author notes:** To whom correspondence should be addressed: Reza Taheri, M.D.

## Abstract

**Objective:** Spina bifida primarily affects people of low and middle socioeconomic status. Herein, we describe the outcome of myelomeningocele surgical management in Iran and predictors of its postoperative complications and mortality.

**Methods:** This retrospective chart review studies the children who underwent surgical management for myelomeningocele in Shiraz, Fars province, Iran, from May 2001 to September 2020. To this end, we investigated mortality and 30-day complications and the factors that determined the operation’s outcome.

**Results:** 256 patients were enrolled. The median age at the operation was roughly eight days (IQR: 7). The most common site of involvement of Myelomeningocele (MMC) was Lumbosacral (86%, n = 204). At the evaluation conducted prior to operation, CSF leaking was observed in 7% (n=16) of the patients. Postoperatively, 5.7% of the patients were expired in the 30 days following the operation (n = 14), while 24% needed readmission (n = 47). The most common complications leading to readmission included wound dehiscence (n = 10, 42%) and wound purulence (n = 6, 25%). No variable was significantly associated with postoperative complication except for the site of the lesion (*p-value* = 0.035) and the presence of the lipid content in the defect (*p-value* = 0.044).

**Conclusions:** Most patients born with MMC are referred for the neurosurgical evaluation following their birth; however, as results show there is much left to be desired compared with the 48h recommended by The Congress of Neurological Surgeons. Here, we concluded that presence of lipid compartment in the lesion and the site of the lesion are the two factor that were associated with the rate of mortality. However, further investigation into preoperative interventions and risk factors to mitigate risk of postoperational complications and mortality is highly encouraged. We highly advocate for the investigation and dissemination of the outcome of the conventional surgical management of MMC in financially restrained areas; because they can show the limitation these settings are confronted with (that are in a way unique to them and different from the resourceful settings) and provide a model for other similar areas with limited suitable care.

## Introduction

Spina bifida is the second major cause of congenital disorders following congenital heart defects and the most common central nervous system malformation compatible with life ^20^.

The most significant type of Spina bifida is myelomeningocele (MMC; open spina bifida), in which the spinal neural tube fails to close properly during embryonic development^4^. Although one of the most common congenital malformations, its underlying mechanism is largely unknown ^4^. MMC has been associated with various disorders, including the Arnold-Chiari II malformation, hydrocephalus, cognitive disability, motor, bowel, and bladder dysfunction, and musculoskeletal abnormalities ^19^. MMC affects the quality of life of the patients during childhood, adolescence, and into adulthood, carrying implications for individuals, families, and society ^4^. Timely detection and complete correction can remarkably minimize the burden and neurological disability ^26^.

Prenatal diagnosis of MMC by ultrasonography enables the timely termination of pregnancy ^4^. In the current convention of treating MMC, two options are available: open and fetal MMC repair. The first step in determination of treatment plan is using MRI or CT scanning to define the nature of the lesion ^18^. The main goal of fetal MMC correction is to improve children’s development and quality of life; however, it is associated with maternal and fetal risks ^1,10^.

Herein, we conducted a study on the outcome of corrective surgery of myelomeningocele in the Iranian population, the factors it relies on, and the complications it causes.

## Methods and Materials

### Patient Population

This retrospective chart review study investigates children <18 years old who underwent initial surgical treatment for myelomeningocele between May 2001 and September 2020 at Namazee hospital, an academic tertiary referral center. Children operated on for any forms of spina bifida other than myelomeningocele during the study window were not be included. Our center has been chosen as a center of referral for surgical correction of MMC by World Health Organization for the Afghans. This has been reflected in the results in the nationality of the patients; however, due to limited accessibility, we had to exclude many of the afghan patients due to limitations in availability of their data in our registry. Data extraction was carried out for the following variables: age at surgery (days), gender, nationality, concomitant hydrocephalus (as determined by need for any operation related to hydrocephalus management in the initial 30 days follow-up window), anatomic site of the lesion, preoperative cerebrospinal fluid leak, presence of lipid compartment in the lesion, difficult dural closure (primary or secondary dural graft), laminectomy, and length of stay in the hospital.

### Statistical Analysis

Statistical analysis was carried out using Fisher’s Exact test or independent-samples t-tests as appropriate. Statistical significance was set a priori at P < 0.05. Statistical analysis was completed using IBM SPSS Statistics version 16 (IBM Corp., Armonk, N.Y., USA).

## Results

**Table 1**.

**Table 1.**
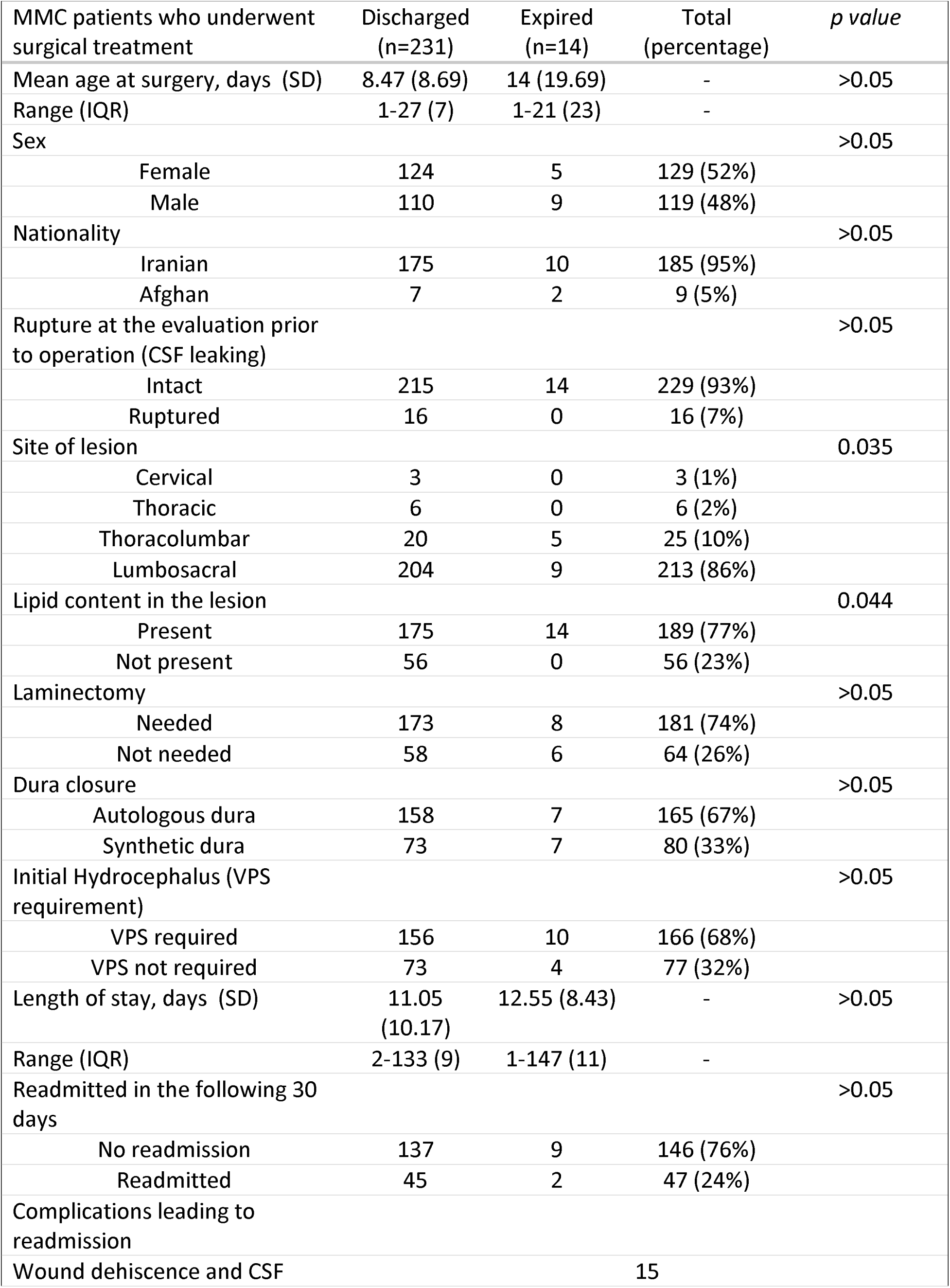
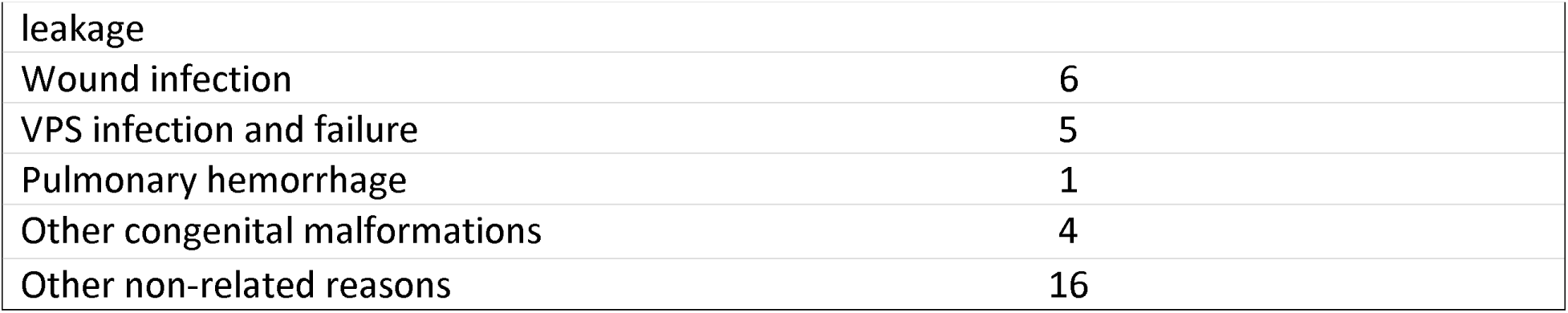
The relationship between patient demographics, pre- and post-operational presentations and mortality in children undergoing surgical treatment of myelomeningocele. IQR: Interquartile Range, VPS: Ventriculoperitoneal Shunt, CSF; Cerebrospinal Fluid

| MMC patients who underwent surgical treatment | Discharged (n=231) | Expired (n=14) | Total (percentage) | *p value* |
| --- | --- | --- | --- | --- |
| Mean age at surgery, days (SD) | 8.47 (8.69) | 14 (19.69) | - | >0.05 |
| Range (IQR) | 1-27 (7) | 1-21 (23) | - |  |
| Sex |  |  |  | >0.05 |
| Female | 124 | 5 | 129 (52%) |  |
| Male | 110 | 9 | 119 (48%) |  |
| Nationality |  |  |  | >0.05 |
| Iranian | 175 | 10 | 185 (95%) |  |
| Afghan | 7 | 2 | 9 (5%) |  |
| Rupture at the evaluation prior to operation (CSF leaking) |  |  |  | >0.05 |
| Intact | 215 | 14 | 229 (93%) |  |
| Ruptured | 16 | 0 | 16 (7%) |  |
| Site of lesion |  |  |  | 0.035 |
| Cervical | 3 | 0 | 3 (1%) |  |
| Thoracic | 6 | 0 | 6 (2%) |  |
| Thoracolumbar | 20 | 5 | 25 (10%) |  |
| Lumbosacral | 204 | 9 | 213 (86%) |  |
| Lipid content in the lesion |  |  |  | 0.044 |
| Present | 175 | 14 | 189 (77%) |  |
| Not present | 56 | 0 | 56 (23%) |  |
| Laminectomy |  |  |  | >0.05 |
| Needed | 173 | 8 | 181 (74%) |  |
| Not needed | 58 | 6 | 64 (26%) |  |
| Dura closure |  |  |  | >0.05 |
| Autologous dura | 158 | 7 | 165 (67%) |  |
| Synthetic dura | 73 | 7 | 80 (33%) |  |
| Initial Hydrocephalus (VPS requirement) |  |  |  | >0.05 |
| VPS required | 156 | 10 | 166 (68%) |  |
| VPS not required | 73 | 4 | 77 (32%) |  |
| Length of stay, days (SD) | 11.05 (10.17) | 12.55 (8.43) | - | >0.05 |
| Range (IQR) | 2-133 (9) | 1-147 (11) | - |  |
| Readmitted in the following 30 days |  |  |  | >0.05 |
| No readmission | 137 | 9 | 146 (76%) |  |
| Readmitted | 45 | 2 | 47 (24%) |  |
| Complications leading to readmission |  |  |  |  |
| Wound dehiscence and CSF |  |  | 15 |  |

|  |  |
| --- | --- |
| leakage |  |
| Wound infection | 6 |
| VPS infection and failure | 5 |
| Pulmonary hemorrhage | 1 |
| Other congenital malformations | 4 |
| Other non-related reasons | 16 |

256 patients were enrolled (Females 52%; n = 129). The mean age at the operation was roughly eight days (IQR= 7). The most common site of involvement of MMC was Lumbosacral (86%, n = 204). At the evaluation conducted prior to operation, CSF leaking was observed in 7% (n = 16) of the patients.

Initial hydrocephalus was observed in 68% (n = 166) of the patients. Postoperatively, 5.7% of patients were expired in the 30 days following the operation (n = 14), while 24% needed a readmission (n = 47). The most common complications leading to readmission included wound dehiscence (n = 10, 42%) and wound purulence (n = 6, 25%). The average length of stay of the patients in the hospital was 11.14 (SD = 10.07).

None of the variables was significantly associated with postoperative complications except for the site of the lesion (*p-value* = 0.035) and the presence of the lipid content in the defect (*p-value* = 0.044). Gender and nationality were not associated with any of the predictors or complications studied (*p value*> 0.05). None of the variables was associated with a higher risk of VentriculoPeritoneal Shunt (VPS) requirement (*p value*> 0.05).

## Discussion

The prevalence of Neural Tube Defects (NTDs) at birth varies considerably depending on the socioeconomic status and the effectiveness of the prenatal screening system. Prevalence of NTDs in the developed countries is estimated at 0.5-0.8/1000 births while in some regions of China, the prevalence was approximately 20 times higher ^8,13^. EUROCAT estimated that the prevalence of ‘spina bifida’ and ‘NTDs’ in stillbirths and pregnancy terminations is 0.51 and 0.94 per 1000 births, respectively ^6^. The mortality rate of spina bifida patients is roughly 1% from age 5 to 30 annually, with lesions at higher levels causing the majority of the mortalies^2,14,24^. Another study pointed out that lesions located on the more rostral functional levels were linked with higher rates of hydrocephalus ^11^. Mortality rates in infants receiving MMC repair range from 7% in low-income countries to 3% in high-income countries such as the United States ^1,12,23^. In the present study, the mortality rate was about 6%, closer to the low-income countries. The Congress of Neurological Surgeons advocates for surgical repair of MMC within 48 hours of birth^17^, which was not accomplished in our setting. Although this recommendation is claimed to improve patient outcomes, it is difficult to achieve for many countries with lower disposable resources. Moreover, this study, due to it retrospective nature, possibly underestimates the rate of mortality because the more severely afflicted patients might die before the admission and the ensuing operation.

Genetics, maternal diminished folate status, maternal obesity, ambient air pollution, smoking, and maternal alcohol use are among the most important risk factor for NTDs ^3,4,9,15,21,27,28^. Studies suggest that routine ultrasonography is sensitive enough to detect more than 90% of spina bifida patients prenatally^4^. However, Joel Haakon Borgstedt-Bakke et al. reported that prenatal detection did not affect mortality^2^. Although ultrasonography is quite effective in the early prenatal diagnosis, poor adherence to prenatal screening in some settings, its limited availability in some others, and refusal by the parents to end the pregnancy in a timely manner lead to the abnormally high incidence of MMC. The family’s willingness to seek medical care, ability to obtain care, and the availability of the appropriate medical care pose numerous challenges for neurosurgeons struggling to provide timely access to medical care for those born with MMC. As Reynolds et al. pointed out, the willingness for seeking care can be affected by the stigma of giving birth to a disabled kid, especially when we consider that most of the cases with MMC are born in family with low socioeconomic status and deprived areas ^23^

Helen J Sims-Williams and colleagues reported the ten-year survival of patients undergoing surgical intervention for MCC to be 55%, with 78% of deaths occurring in the first five years. The majority of the deaths were not directly related to MMC; conversely, infection and failure of the caregivers to take proper care constituted the leading causes of deaths. Compared with the operation conducted within the first 48 hours of birth, delayed surgery at 15-29 days was associated with higher mortality ^25^. In the current study, no relationship between early and delayed intervention was observed.

Kim et al. determined that 80% (68% in the current study) of the myelomeningocele patients registered at National Spina Bifida Patient Registry (NSBPR) were also treated for hydrocephalus^11^. Another study pointed out that there was no statistically significant difference between lesion functional levels with respect to the patients’ ages at the timing of the first shunt insertion ^22^. None of the factors studied here were associated with VPS requirement.

Rebecca A. Reynolds et al. showed that 31% of the patients experienced a complication compared to 24% in the current study. The most common complications in MMC patients included wound dehiscence and wound purulence^23^. In patients undergoing shunt insertion, the overall rates of shunt infection and shunt malfunction were 16.5 and 39.4%, respectively^22^.

Thus far, most of the body of research concerned with MMC comes from the US. Despite the high prevalence of MMC in undeveloped countries, our knowledge regarding MMC in those settings is exceedingly limited. We previously aspired to demonstrate the requisition to localize the data, in that the confrontation strategy differs in various settings.; because the incidence, the rate of success of interventions, the associated complications, etc. are dissimilar^16^. In the light of such studies, issues that have not been confronted in a health care system with relatively more resources like that of the US come to light and we begin to realize that issues as simple as managing the urological complications of MMC in the long-term become huge and exhausting due to lack of robust follow-up and referral systems.

This study may not have captured a minority of the patients who died at home because some of them were lost to follow-up. Moreover, there possibly is a sampling bias. This retrospective study merely includes the patients who underwent an operation, which possibly selects for healthier infants with more preferable prognoses as determined by the medical team. In spite of these limitations, the present study serves as a platform for further research that will better inform us on the prognosis and risk factors, and provides a chance to investigate and intervene on modifiable risk factors examined in this study in efforts to improve the patient outcomes.

## Data Availability

All data generated or analyzed during this study are included in the final published article.

## Competing interests

The authors have no relevant financial or non-financial interests to disclose. Availability of data and material

All data generated or analyzed during this study are included in the final published article.

## Authors’ contributions

This study was designed by Mohammad Sadegh Masoudi, Sina Zoghi, and Reza Taheri. The data was collected by Maryam Feili, Mohammad Amin Mosayebi, and Mohammad Amin Afifi. Data curation and analysis were conducted by Sina Zoghi. The final manuscript was written and edited by Sina Zoghi, Maryam Feili, and Afrooz Feili. All contributing authors approved the final manuscript.

## Ethical approval and consent to participants

Ethical approval was waived in view of the retrospective nature of the study and all the procedures being performed were part of the routine care. Ethical consent for participation and publication was collected when the operations were conducted.

## Reference

1. Adzick NS, Thom EA, Spong CY, Brock III JW, Burrows PK, Johnson MP, et al: A randomized trial of prenatal versus postnatal repair of myelomeningocele. N Engl J Med 364:993–1004, 2011

2. Borgstedt-Bakke JH, Fenger-Grøn M, Rasmussen MM: Correlation of mortality with lesion level in patients with myelomeningocele: a population-based study. J Neurosurg Pediatr PED 19:227–231, 2016 Available: https://thejns.org/pediatrics/view/journals/j-neurosurg-pediatr/19/2/article-p227.xml.

3. Carter CO, Evans K: Spina bifida and anencephalus in greater London. J Med Genet 10:209–234, 1973 Available: https://pubmed.ncbi.nlm.nih.gov/4590246.

4. Copp AJ, Adzick NS, Chitty LS, Fletcher JM, Holmbeck GN, Shaw GM: Spina bifida. Nat Rev Dis Prim 1:15007, 2015 Available: https://pubmed.ncbi.nlm.nih.gov/27189655.

5. Dicianno BE, Karmarkar A, Houtrow A, Crytzer TM, Cushanick KM, McCoy A, et al: Factors Associated with Mobility Outcomes in a National Spina Bifida Patient Registry. Am J Phys Med Rehabil 94:1015–1025, 2015 Available: https://pubmed.ncbi.nlm.nih.gov/26488146.

6. Dolk H, Loane M, Garne E: The prevalence of congenital anomalies in Europe. Rare Dis Epidemiol:349–364, 2010

7. Farmer DL, Thom EA, Brock III JW, Burrows PK, Johnson MP, Howell LJ, et al: The Management of Myelomeningocele Study: full cohort 30-month pediatric outcomes. Am J Obstet Gynecol 218:256–e1, 2018

8. Flynt JW JW F: International clearinghouse for birth defects monitoring systems. 1979

9. Grewal J, Carmichael SL, Ma C, Lammer EJ, Shaw GM: Maternal periconceptional smoking and alcohol consumption and risk for select congenital anomalies. Birth Defects Res A Clin Mol Teratol 82:519–526, 2008 Available: https://pubmed.ncbi.nlm.nih.gov/18481814.

10. Kabagambe SK, Jensen GW, Chen YJ, Vanover MA, Farmer DL: Fetal Surgery for Myelomeningocele: A Systematic Review and Meta-Analysis of Outcomes in Fetoscopic versus Open Repair. Fetal Diagn Ther 43:161–174, 2018 Available: https://www.karger.com/DOI/10.1159/000479505.

11. Kim I, Hopson B, Aban I, Rizk EB, Dias MS, Bowman R, et al: Treated hydrocephalus in individuals with myelomeningocele in the National Spina Bifida Patient Registry. J Neurosurg Pediatr PED 22:646–651, 2018 Available: https://thejns.org/pediatrics/view/journals/j-neurosurg-pediatr/22/6/article-p646.xml.

12. Leidinger A, Piquer J, Kim EE, Nahonda H, Qureshi MM, Young PH: Experience in the early surgical management of myelomeningocele in Zanzibar. World Neurosurg 121:e493–e499, 2019

13. Li Z, Ren A, Zhang L, Ye R, Li S, Zheng J, et al: Extremely high prevalence of neural tube defects in a 4-county area in Shanxi Province, China. Birth Defects Res Part A Clin Mol Teratol 76:237–240, 2006

14. Liptak GS, Kennedy JA, Dosa NP: Youth with spina bifida and transitions: health and social participation in a nationally represented sample. J Pediatr 157:584–588, 2010

15. Lupo PJ, Symanski E, Waller DK, Chan W, Langlois PH, Canfield MA, et al: Maternal exposure to ambient levels of benzene and neural tube defects among offspring: Texas, 1999-2004. Environ Health Perspect 119:397–402, 2011 Available: https://pubmed.ncbi.nlm.nih.gov/20923742.

16. Masoudi MS, Taheri R, Zoghi S: Predictive Factors for Post-Operative Tracheostomy Requirement in Children Undergoing Surgical Resection of Medulloblastoma. World Neurosurg: 2021

17. Mazzola CA, Assassi N, Baird LC, Bauer DF, Beier AD, Blount JP, et al: Congress of Neurological Surgeons Systematic Review and Evidence-Based Guidelines for Pediatric Myelomeningocele: Executive Summary. Neurosurgery 85:299–301, 2019 Available: https://doi.org/10.1093/neuros/nyz261.

18. McComb JG: Spinal and cranial neural tube defects. Semin Pediatr Neurol 4:156–166, 1997

19. McDowell MM, Blatt JE, Deibert CP, Zwagerman NT, Tempel ZJ, Greene S: Predictors of mortality in children with myelomeningocele and symptomatic Chiari type II malformation. J Neurosurg Pediatr 21:587–596, 2018

20. Mohd-Zin SW, Marwan AI, Abou Chaar MK, Ahmad-Annuar A, Abdul-Aziz NM: Spina Bifida: Pathogenesis, Mechanisms, and Genes in Mice and Humans. Scientifica (Cairo) 2017:5364827, 2017

21. Padula AM, Tager IB, Carmichael SL, Hammond SK, Lurmann F, Shaw GM: The association of ambient air pollution and traffic exposures with selected congenital anomalies in the San Joaquin Valley of California. Am J Epidemiol 177:1074–1085, 2013 Available: https://pubmed.ncbi.nlm.nih.gov/23538941.

22. Radmanesh F, Nejat F, Khashab M El, Ghodsi SM, Ardebili HE: Shunt complications in children with myelomeningocele: effect of timing of shunt placement. J Neurosurg Pediatr PED 3:516–520, 2009 Available: https://thejns.org/pediatrics/view/journals/j-neurosurg-pediatr/3/6/article-p516.xml.

23. Reynolds RA, Bhebhe A, Garcia RM, Chen H, Bonfield CM, Lam S, et al: Surgical Outcomes after Myelomeningocele Repair in Lusaka, Zambia. World Neurosurg 145:e332–e339, 2021

24. Sawyer SM, Collins N, Bryan D, Brown D, Hope MA, Bowes G: Young people with spina bifida: transfer from paediatric to adult health care. J Paediatr Child Health 34:414–417, 1998

25. Sims-Williams HJ, Sims-Williams HP, Kabachelor EM, Fotheringham J, Warf BC: Ten-year survival of Ugandan infants after myelomeningocele closure. J Neurosurg Pediatr 19:70–76, 2017

26. Venkataramana NK: Spinal dysraphism. J Pediatr Neurosci 6:S31–40, 2011

27. Wahbeh F, Manyama M: The role of Vitamin B12 and genetic risk factors in the etiology of neural tube defects: A systematic review. Int J Dev Neurosci n/a: 2021 Available: https://doi.org/10.1002/jdn.10113.

28. Waller DK, Shaw GM, Rasmussen SA, Hobbs CA, Canfield MA, Siega-Riz A-M, et al: Prepregnancy Obesity as a Risk Factor for Structural Birth Defects. Arch Pediatr Adolesc Med 161:745–750, 2007 Available: https://doi.org/10.1001/archpedi.161.8.745.

